# Modeling fractional order dynamics of Syphilis via Mittag-Leffler law

**DOI:** 10.1101/2021.02.05.21251119

**Authors:** E. Bonyah, C.W. Chukwu, M.L. Juga, Fatmawati

**Author notes:** Corresponding author: Email address (C.W. Chukwu).

## Abstract

Syphilis is one the most dangerous sexually transmitted disease which is common in the world. In this work, a mathematical model is formulated with an emphasis on treatment. The reproduction number which presents information on the spread of the disease is determined. The model’s steady states are established, and the disease free state’s local and global stability are studied. The existence and uniqueness of solutions for both Caputo-Fabrizio and Atangana-Baleanu derivative in the Caputo sense are established. Numerical simulations were carried out to support the analytical solution, which indicates that the fractional order derivatives influence the dynamics of the spread of the Syphilis in the community.

*2010 MSC*: 00-01, 99-00

## 1. Introduction

Syphilis has remained a persistent human health threat in both developed and developing countries [1, 2]. It is a sexually transmitted infection caused by the bacterium Treponema Palladum which is said to infect approximately 12 million people around the globe yearly. Syphilis is transmitted from person to person by direct contact with a syphilitic sore, known as a chancre. Chancres can occur on or around the external genitals, in the vagina, around the anus, rectum, or in or around the mouth. Transmission of Syphilis can occur during vaginal, anal, or oral sex [3]. It has several symptoms, most of which are also common to other diseases. If not properly treated, it can progress from primary to secondary and finally to the disease’s tertiary stage.

Syphilis infection is characterised by an ulcerative chancre signaling the beginning of the primary stage of the disease [4]. After exposure and infection, the primary incubation period is about 25 days, although available data suggest that this period can be between 3 and 4 weeks, [5] (3–6 weeks according to the CDC, [3]). If not treated, the disease progresses to the secondary stage with symptoms like skin rashes and mucous membrane lesions,[4] and an incubation period of about 46 days, [5]. Following the secondary symptoms, the infection progresses to the tertiary and latent stage where the disease remains in the body and can reappear or even damage internal organs or lead to death, [4, 3].

Syphilis can be treated with antibiotics such as penicillin, [4, 6]. After treatment and recovery from the infection, individuals may develop transitory immunity to reinfection before becoming susceptible again, [4], although it seems immunity depends on the stage of the disease at which treatment was implemented, [4, 7, 5].

One of the early triumphs of mathematical epidemiology was a formulation to predict the dynamics of a disease. Mathematical models use some basic assumptions and mathematics to find parameters for various infectious diseases and use those parameters to calculate the effects of possible interventions, [8, 9, 10, 11, 12, 13, 14]. A lot of these models have been developed to study the dynamics of Syphilis transmission. One of the models included the different stages of the disease and treatment [5], while another used 210 differential equations to model heterosexual Syphilis transmission in East Vancouver. Here, they combined the later stages of Syphilis but partitioned the population into multiple groups based on sex, sexual activity and age, [15]. In a more recent study, Milner and Zhao [16] presented an ODE model based on partial immunity and vaccination (assuming a successful vaccine is developed), and showed that there exists backward bifurcation for some parameter values. Despite the diversity Various methodologies in the existing Syphilis models are integer-order models that give inaccurate predictions due to their lack of memory effect.

In the last few years, Fractional calculus has gained a lot of popularity because of its application in many areas and its ability to consider the memory effect, which is a natural occurrence in several biological models. Riemann and Louiville first proposed a fractional derivative with singular kernel. Next Caputo and Fabrizio in [17] presented a new definition of fractional derivative without singular kernel, which proved to be fair and applied by many researchers. A few years ago, Atangana and Baleanu developed a new operator which is based on the generalized Mittag-Leffler function where the kernel is non-singular and non-local, [18]. Several non-integer order models in the sense of Caputo-Fabrizio and in the sense of Atangana-Baleanu have been developed [19, 20, 10, 21, 22]. However, none of them compared the results of syphilis transmission dynamics in the sense of Caputo-Fabrizio (CF), with that in the Atangana -Baleanu sense. In this paper, we develop a mathematical model to study the dynamics of Syphilis transmission via Mittag-Leffler law and compare the results obtained via the Caputo-Fabrizio derivative.

The first Section is a brief introduction, followed by the model formulation in Section 2. We give the model analysis in Section 3 and introduce the CF operator in Section 4, together with the preliminaries and model properties. Section 5 presents a numerical scheme for the ABC model, while Section 6 presents the numerical simulations followed by the conclusion.

## 2. Syphilis model formulation

In this section, we describe the transmission dynamics of Syphilis disease. The model comprises of five compartments, which are the Susceptible individuals (*S*(*t*)), Exposed individuals (*E*(*t*)), individuals at the early stage of Syphilis infection (*I*(*t*)), individuals at the late stage of Syphilis infection (*L*(*t*)), and individuals treated of Syphilis infection (*T* (*t*)). The Susceptible individuals are recruited into the population at a rate Π. These individuals come in contact with those in *I*(*t*) and *L*(*t*) and contract the disease at a rate *λ*, where

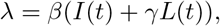

and *β* is the probability that a contact between a susceptible individual and an infectious individual will result to an infection. After being exposed to the bacteria Treponema pallidum, they progress to the class *I*(*t*) at a rate *ρ*_1_. Furthermore, the individuals in *I*(*t*) can either progress to the class *L*(*t*) at a rate *ρ*_2_ or recover after treatment from the Syphilis infection at rate *ν*_1_. On the other hand, the treated individuals can also become exposed again to Syphilis upon interaction with individuals in *I*(*t*) or *L*(*t*) at a rate *ωλ*, while those at the later stage of the infection can also be treated at a rate *ν*_2_. We assume that individuals in all the compartments have a natural mortality rate of *µ*. The flow diagram for the model is shown in Figure 1. The model diagram in Figure 1 together with the assumptions gives rise to the following system of equations:

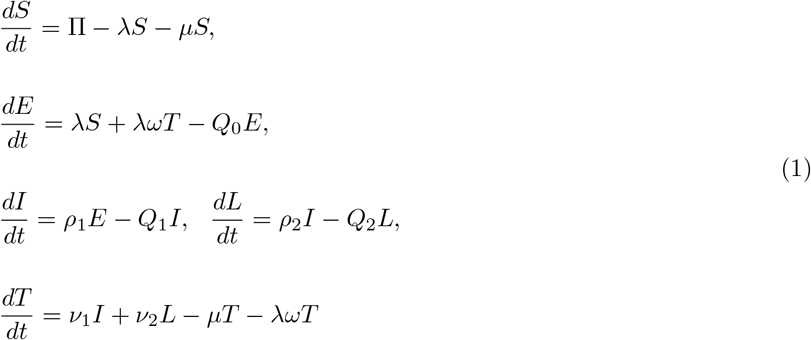

where *Q*_0_ = (*µ* + *ρ*_1_), *Q*_1_ = (*ρ*_2_ + *ν*_1_ + *µ*) and *Q*_2_ = (*ν*_2_ + *δ* + *µ*), with the following initial conditions

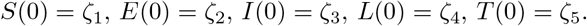

**Figure 1:**
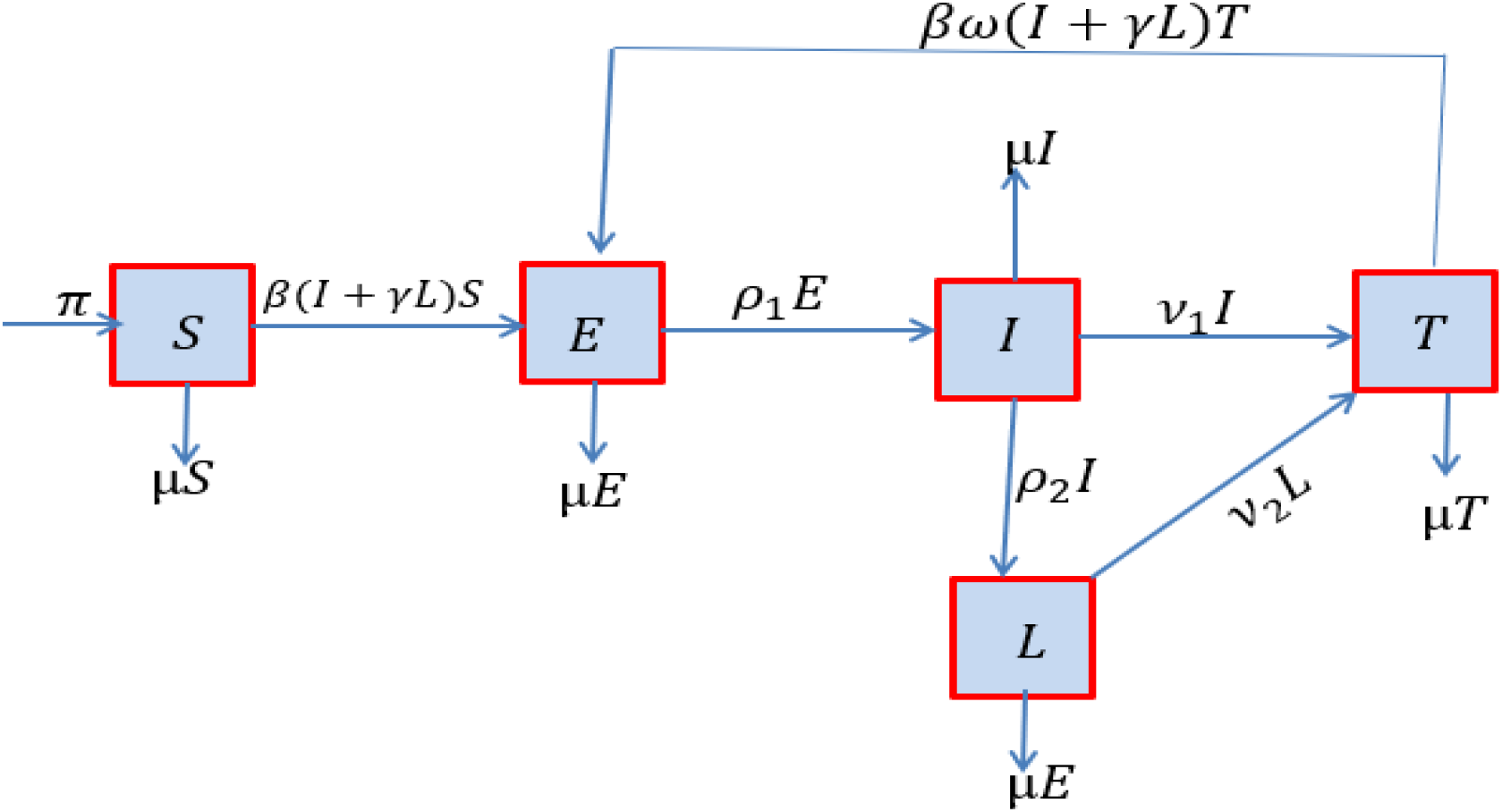
Syphilis model flow diagram.

## 3. Syphilis model with Caputo-Fabrizio

The Syphilis model with Caputo-Fabrizio (CF) derivative is given by

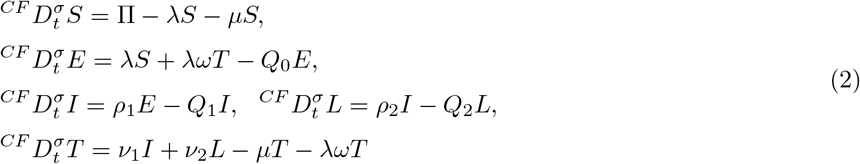

with *σ* being the fractional order 0 < *σ* < 1 subject to the following initial conditions

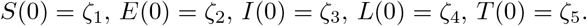

### 3.1. Basic preliminaries

Here, we give some of the mathematical preliminaries in the form of theorems, which we shall apply to prove the positivity and uniqueness and positivity of Syphilis model with Caputo-Fabrizio (2) as defined in [23, 24] respectively. The definitions are stated as follows

#### Definition 1.

Assume *φ*(*t*) ∈ *ℋ* ^1^(*ℓ*_1_, *ℓ*_2_), for *ℓ*_2_ > *ℓ*_1_, *τ* ∈ [0, 1]. The CF fractional operator is given as

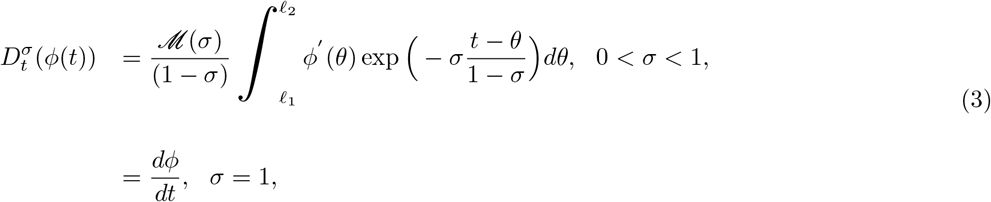

where *ℳ* (*σ*) satisfies the condition *ℳ* (0) = *ℳ* (1) = 1.

#### Definition 2.

The integral operator of fractional order corresponding to the CF fractional derivative is stated as follows

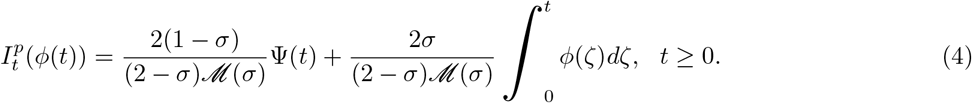

#### Definition 3.

The Laplace transform of 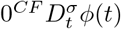 is represented as follows

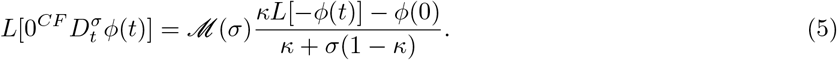

### 3.2. Positivity of solutions of Syphilis model with CF

We prove the positivity of the system using the following theorem.

#### Theorem 1.

Given the initial conditions *S*(0) > 0, *E*(0) ≥ 0, *I*(0) ≥ 0, *T* (0) ≥ 0, *L*_*b*_(0) ≥ 0 for all *t* ≥ 0, we show that the set 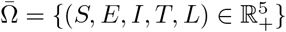 attracts all positive solutions of the fractional order system (2)

We use Lemma 1 to prove Theorem 1.

#### Lemma 1.

Suppose *f* (*t*) ∈ *C*[*a, b*] and 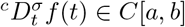 for all 0 < *σ* ≤ 1, then we have 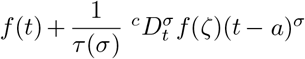, where *a* ≤ *ζ* ≤ *t*, for all *t* ∈ (*a, b*] [25].

Following Lemma 1, we give obtain the following remark.

#### Remark 2.

Assume that *K*(*x*) ∈ *C*[*a, b*] and 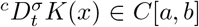 for 0 < *σ* ≤ 1. It follows from Lemma 1 that if 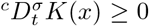, for all *x* ∈ (*a, b*), then *K*(*x*) is non decreasing and if 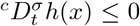 for all *x* ∈ (*a, b*), then *K*(*x*) is non increasing.

We now prove Theorem 1.

*Proof*. Using Lemma 1 and Remark 2 we show that the Syphilis model with CF has exist and has a unique solutions. Here, we prove that 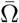 is positively invariant for each hyperplane bonding, the positive orthnant of the vector field points in 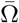. Therefore, model system (2) becomes

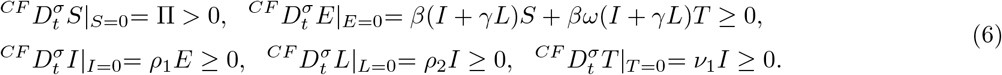

Thus, equation (2) is positively invariant and all its solutions are attracting and positive in 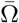 for *t* ≥ 0.

### 3.3. Existence and uniquness of solutions of the CF model

This subsection is devoted to proving the existence and uniqueness of the solution for model (2) by applying the integral operator as defined in Losada and Nieto [24] which yields:

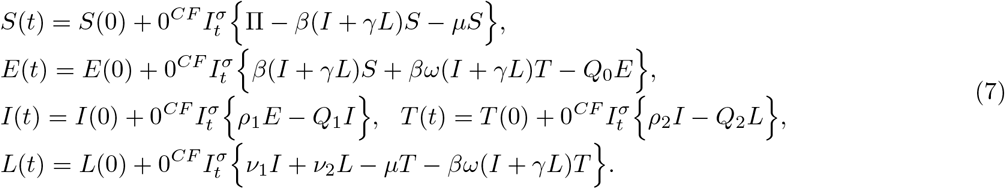

Using the same notation as in [24], equation (7) becomes

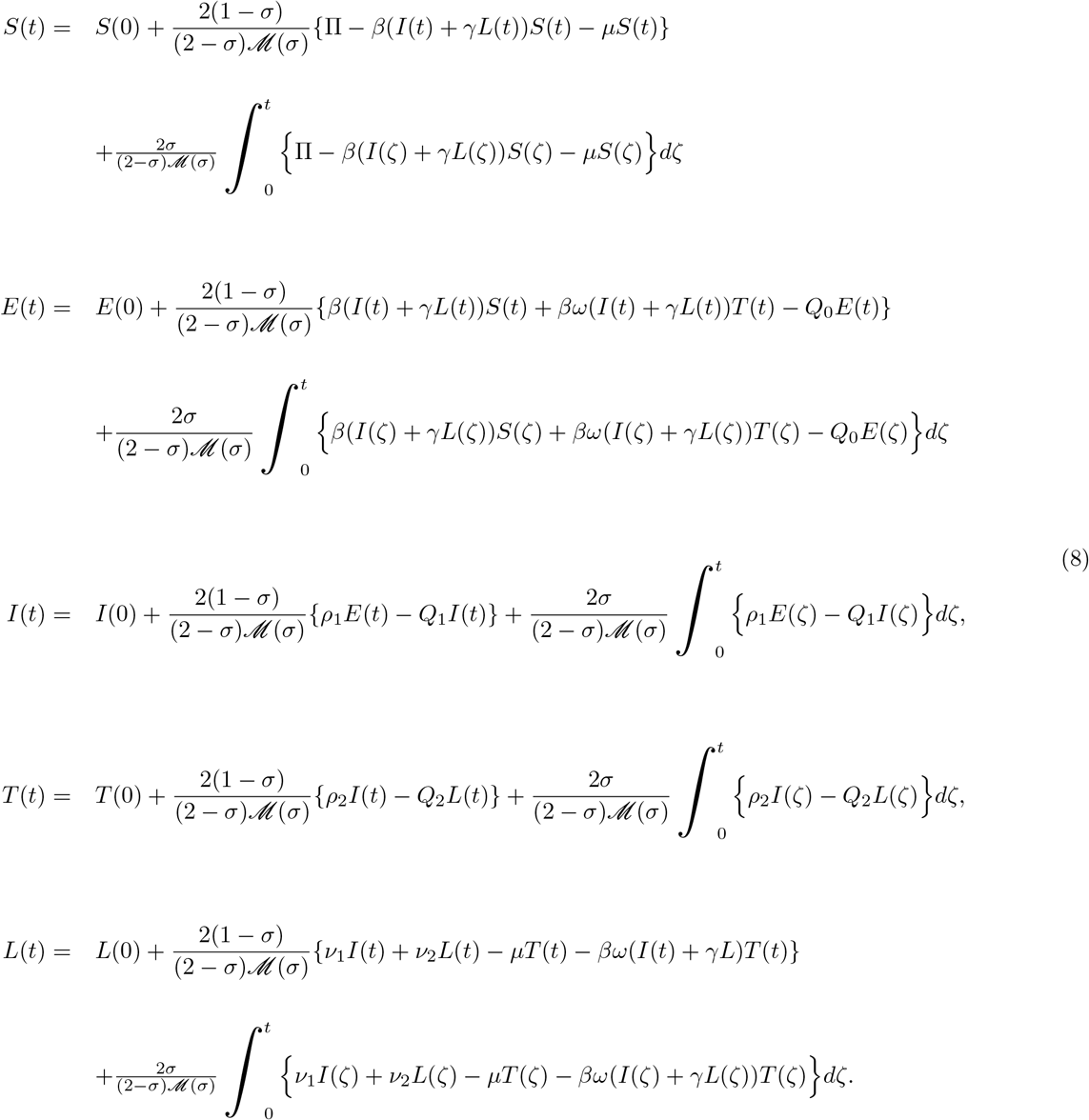

Without loss of generality and for simplification of notations, we shall denote

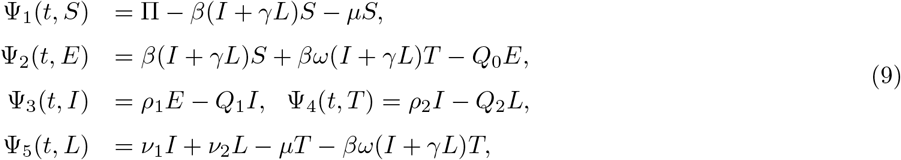

#### Theorem 3.

Each kernel (Ψ_1_, Ψ_2_, Ψ_3_, Ψ_4_, Ψ_5_, Ψ_6_, Ψ_7_) satisfy the Lipschitz condition and contraction if and only if the following inequality holds.

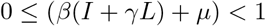

*Proof*. Considering the function *S* and *S*_1_ we have that

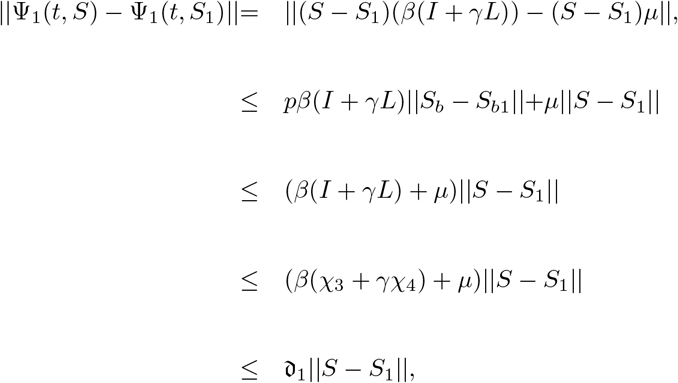

where 𝔡_1_ = *pβ*_*b*_*c*_3_ + *µ*_*b*_, ||*S*(*t*)||≤ *c*_1_, ||*E*(*t*)||≤ *c*_2_, ||*I*(*t*)||≤ *c*_3_, ||*L*(*t*)||≤ *c*_4_ and ||*T* (*t*)||≤ *c*_5_ are all bounded functions. Hence,

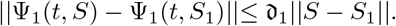

Thus, Ψ_1_ satisfies the Lipschitz condition if 0 ≤ (*β*(*I* + *γL*)*µ*) < 1, and is also a contraction.

Using a similar approach, we can show that (Ψ_2_(*t, E*), Ψ_3_(*t, I*), Ψ_4_(*t, T*), Ψ_5_(*t, L*)) satisfy the Lipschitz conditions.

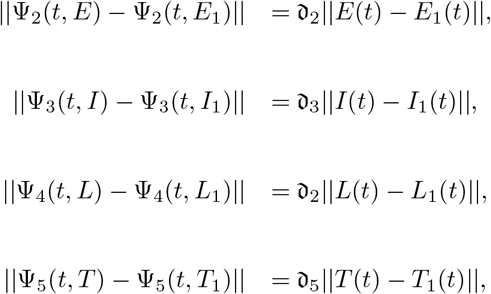

where 𝔡_2_ = *Q*_0_*χ*_2_, 𝔡_3_ = *Q*_1_*χ*_3_, 𝔡_4_ = *Q*_2_*χ*_4_ and 𝔡_5_ = *µχ*_5_ − *βω*(*χ*_3_ + *γχ*_4_)*χ*_5_.

We reduce (9) using the same notation as in equation (4) and obtain:

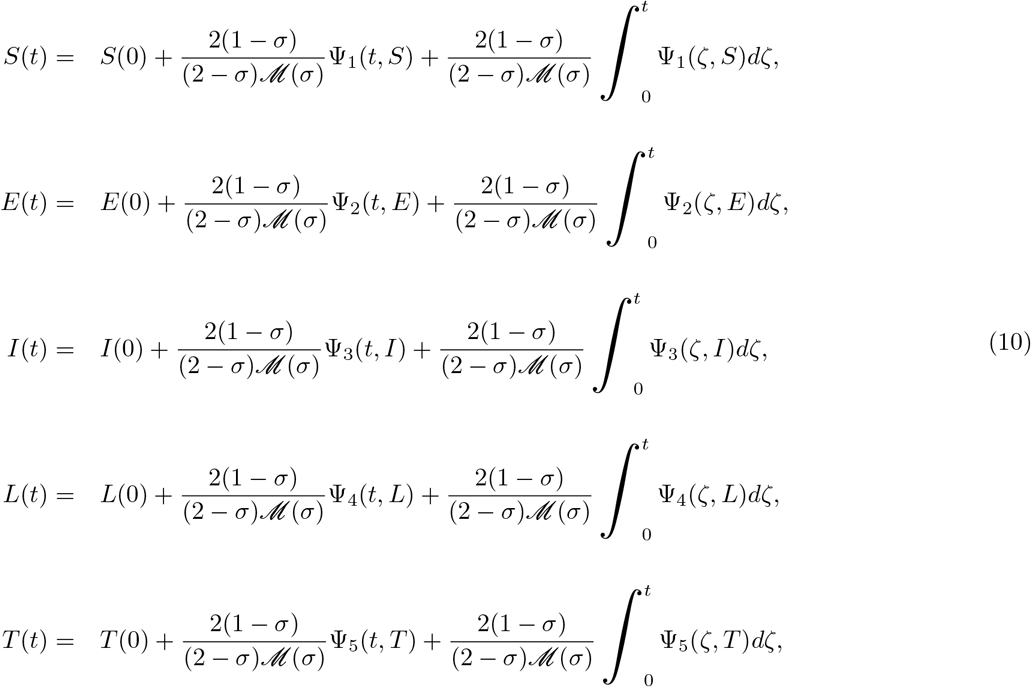

Suppose we define the iterative recursive forms below

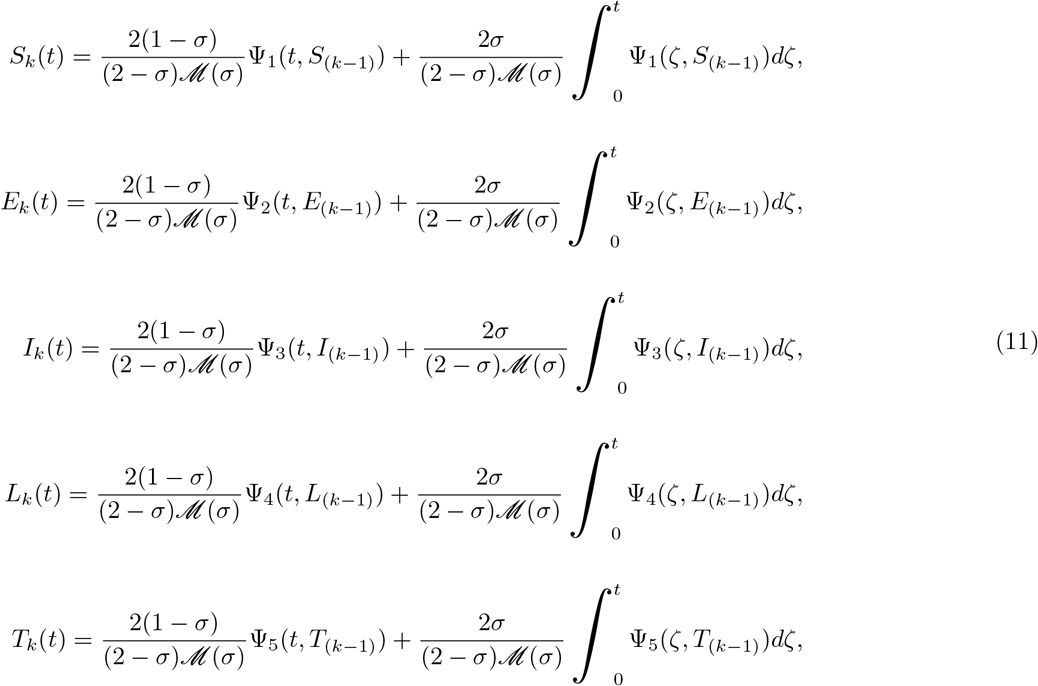

with the initial conditions

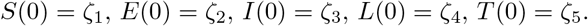

Next, we find the difference between the each successive and obtain the following result:

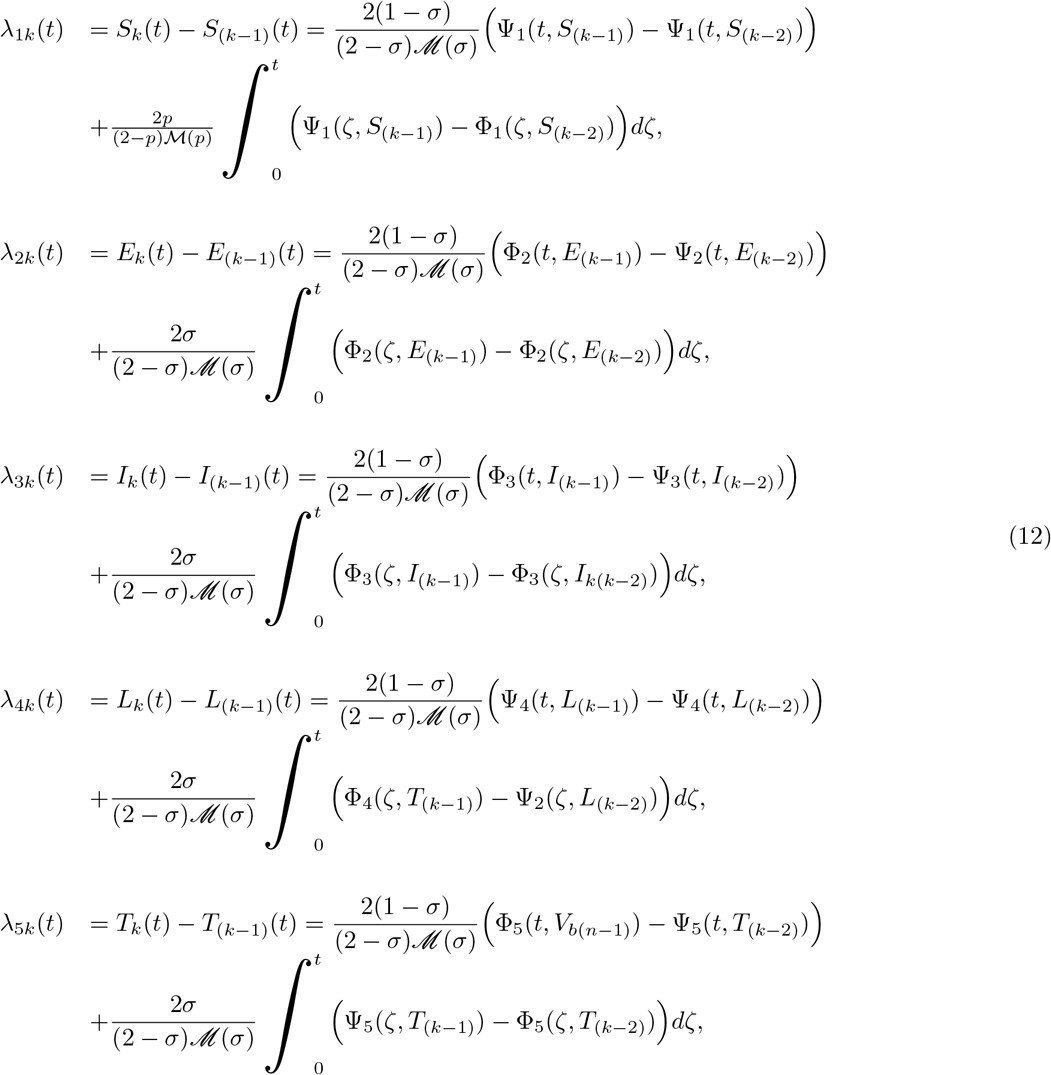

in which

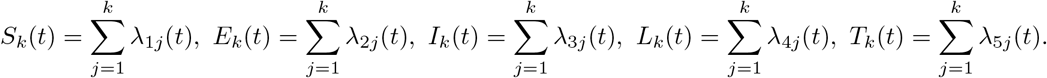

We thus have the following results

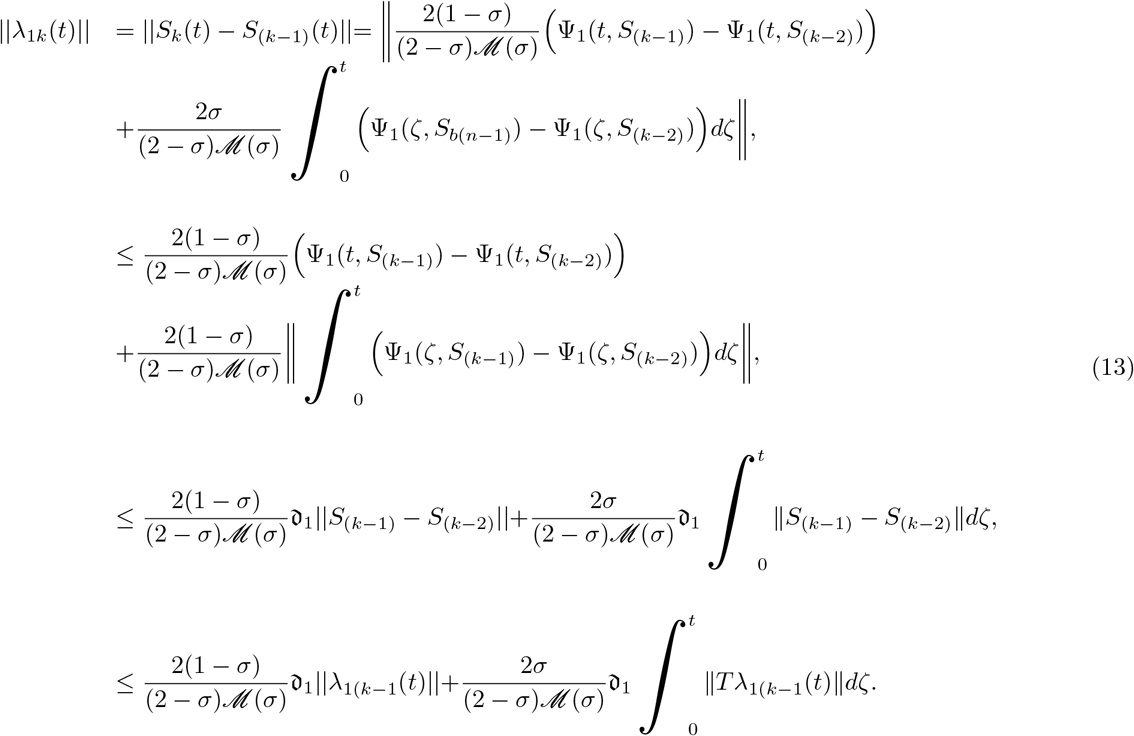

Using a similar approach, we can show that

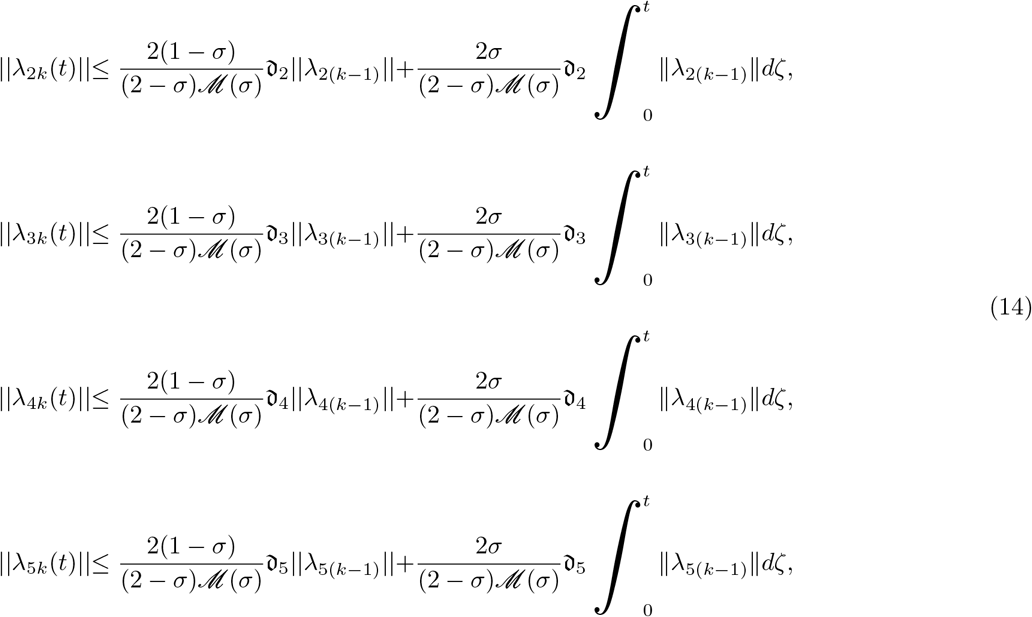

#### Theorem 4.

The Syphilis Caputo-Fabrizio model (2) has a unique solution if

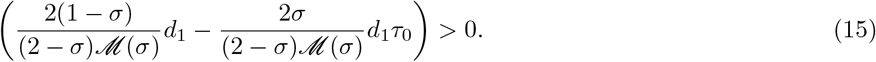

Let equations (13) and (14) bounded functions, we verify that the kernels (Ψ_1_, Ψ_2_, Ψ_3_, Ψ_4_, Ψ_5_, Ψ_6_, Ψ_7_) holds Lipschitz conditions and applying the recursive method and Theorem 4 we obtain

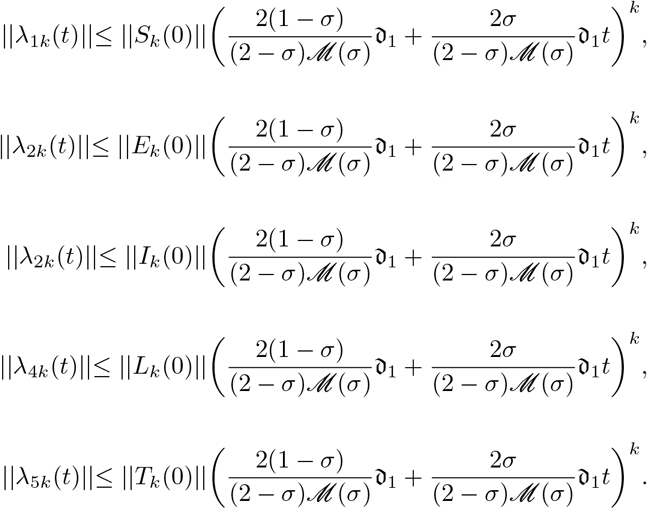

Therefore, we have that equation (15) exists and is a smooth function, continuous and true for any value of *k*.

Further, suppose the fractional model has solutions denoted by *S*^*\**^(*t*), *E*^*\**^(*t*), *I*^*\**^(*t*), *L*^*\**^(*t*) and *L*^*\**^(*t*). we show that there exists a solution of the Syphilis CF fractional model (2) as follows:

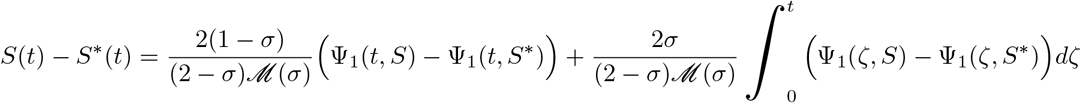

and

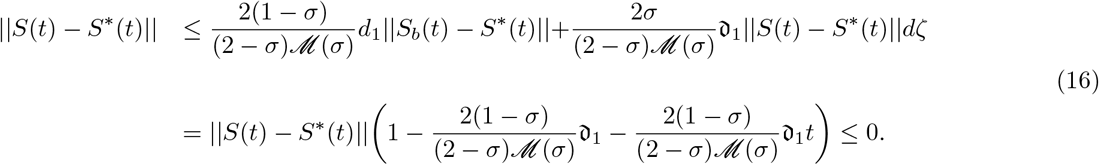

#### Theorem 5.

The model (2) has a unique solution if

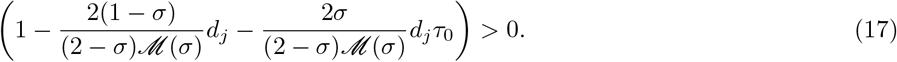

Using results from (16), we have that

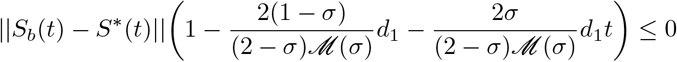

and also from (17) we obtain

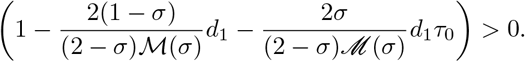

Therefore, ||*S*(*t*) − *S*^*\**^(*t*)||= 0, which implies that *S*(*t*) = *S*^*\**^(*t*). Similarly, we can also establish that *E*(*t*) = *E*^*\**^(*t*), *I*(*t*) = *I*^*\**^(*t*), *L*(*t*) = *L*^*\**^(*t*), *T* (*t*) = *T* ^*\**^(*t*).

## 4. CF model analysis

### 4.1. Model steady states and reproduction number

In this subsection, we present the steady states of model (1) as well as their stabilities. Model (2) has a disease free state (DFE) denoted by *ε*^0^ whenever *E*^*\**^ = *I*^*\**^ = *I*^*\**^ = *L*^*\**^ = *L*^*\**^ = 0, which gives

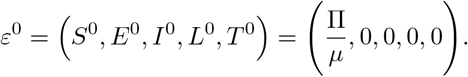

The endemic equilibrium state denoted by *ε*^1^ is given by

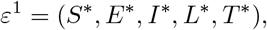

where

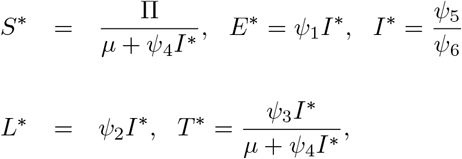

and 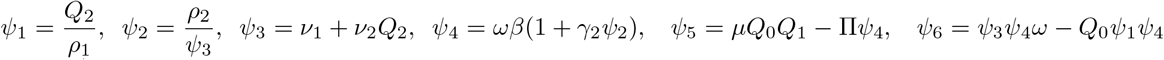. Note that *ε*^1^ exists if and only if Ψ_5_ ≥ 0 and Ψ_6_ > 0.

Next, we determine the basic reproduction number denoted by *ℛ*_*S*_. The parameter *ℛ*_*S*_ in this model is defined as the average number of new infections generated by an infectious individual in an early or late stage of the disease through direct contact with a Syphilis sore in a purely susceptible population. We use the method in [26] to find *ℛ*_*S*_ as follows. Given the Jacobian matrices

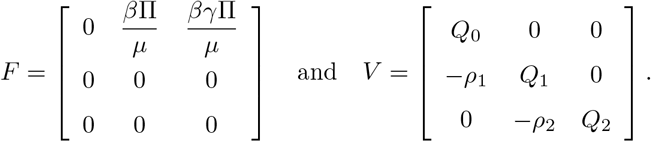

of the new infections and transfer matrices of the CF model (2) (where *F* and *V* are evaluated at the DFE respectively),

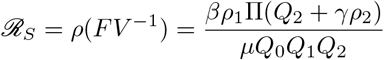

where *FV* ^−1^ is the spectral radius of the next generation matrix, that is *FV* ^−1^.

### 4.2. Local stability of the DFE

#### Theorem 6.

Let *d*_1_,*d*_2_ ∈ ℋ such that gcd(*d*_1_, *d*_2_) = 1 and 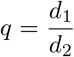. If *M* = *d*_2_, then, the DFE of the system (2) is locally asymptotically stable (LAS) if 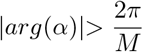 for all roots of *α* of the characteristics equation (18) of the matrix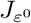,

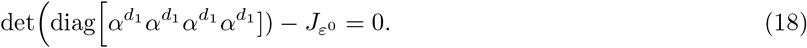

*Proof*. The Jacobian matrix of the system (2) evaluated at the DFE is given by

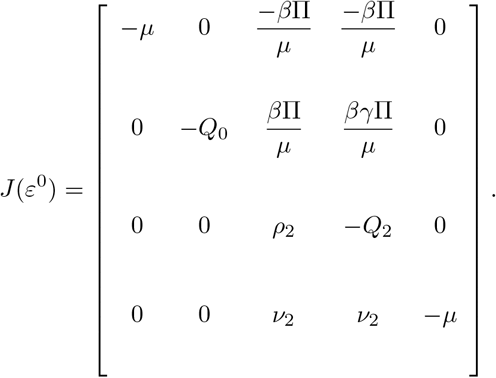

the characteristics equation associated with *J*(*ε*^0^) is

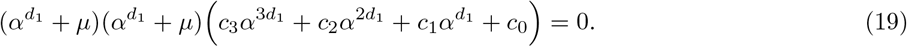

where *c*_0_ = *µQ*_0_*Q*_1_*Q*_2_(*ℛ*_*S*_ − 1), *c*_1_ = Π*βρ*_1_ − *µ*(*Q*_0_*Q*_1_ + *Q*_0_*Q*_2_ + *Q*_1_*Q*_2_), *c*_2_ = −*µ*(*Q*_0_ + *Q*_1_ + *Q*_2_) and *c*_3_ = −*µ*. The arguments of the roots of the equation (*λ*^*d*^1 + *µ*) = 0, (*λ*^*d*^1 + *µ*) = 0 are

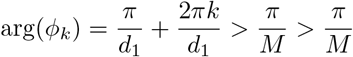

where *k* = 0, 1, …, (*p* − 1). Thus, the first two roots of (19) are −*µ*, − *µ* and the others are calculated from

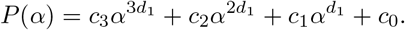

The coefficients of *c*_3_ and *c*_2_ are clearly negative when *ℛ*_*S*_ < 1, *c*_0_ is negative and *c*_1_ is also negative because

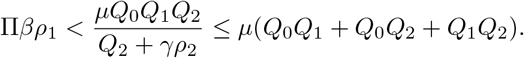

Therefore, when *ℛ*_*S*_ < 1, there is no sign change in the coefficients of *P* (*α*). Thus, by Descartes’s rules of signs, all the roots of *P* (*α*) are either negative or have negative real parts. Hence, the necessary condition for the root of the characteristics equation 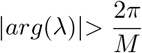 is fulfilled. The DFE is thus, LAS for *ℛ*_*S*_ < 1.

### 4.3. Global stability of the DFE

#### Theorem 7.

The DFE is globally stable for *ℛ*_*S*_ < 1 and unstable otherwise.

*Proof*. We use the *C*^1^ Lyapnouv function

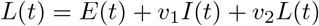

which is made up of the compartments that directly contribute to disease transmission and *v*_1_ and *v*_2_ are positive constants. The CF derivative of *L*(*t*) is

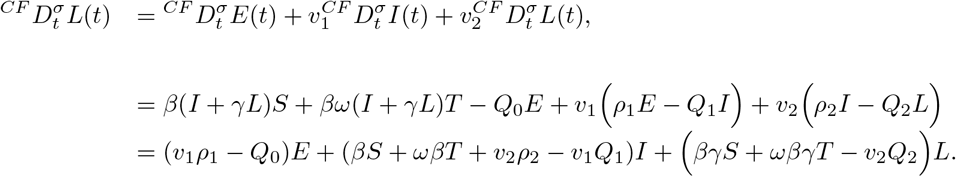

At DFE, 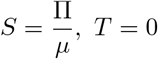. Therefore, *L*(*t*) satisfies the inequality

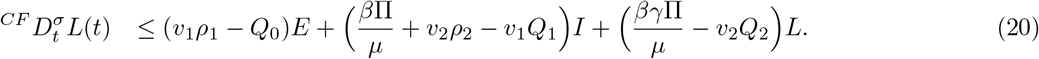

Equating the coefficients of *I* and *L* to zero we obtain the values of *v*_1_ and *v*_2_ as follows

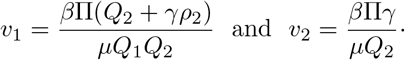

Substituting the constants *v*_1_ and *v*_2_ into the inequality (20) we have that *Q*_0_(*ℛ*_*S*_ − 1)*E* ≤ 0. Whenever *ℛ*_*S*_ ≤ 1, 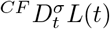 is negative with equality when *ℛ*_*S*_ = 1. Thus, by LaSalle’s invariant principle [27] the DFE is globally stable in the invariant region and unstable otherwise.

### 4.4. Numerical scheme for the CF model

In this subsection, we derive the numerical scheme of the syphilis model with CF using the method of using two-step fractional Adams-Bashforth technique for the CF fractional derivative as in [28] and [29]. We re-write the equation (2) as follows

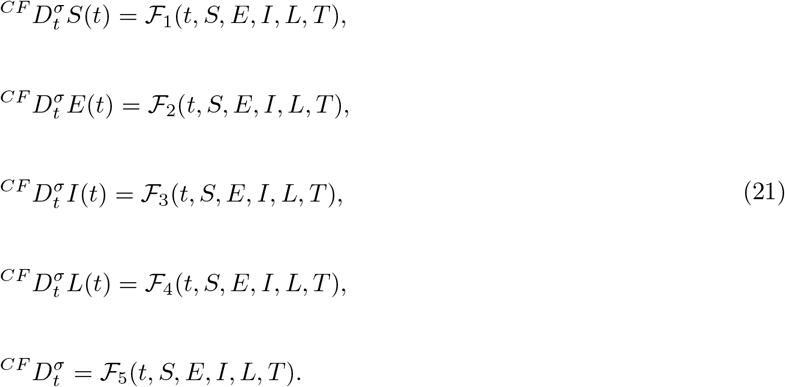

Using the fundamental theorem of fractional calculus, the first equation of system (21) is converted to

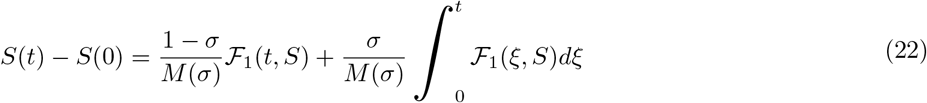

For *t* = *t*_*n*+1_, *n* = 0, 1, 2, …, we have *S*(*t*_*n*+1_) we obtain

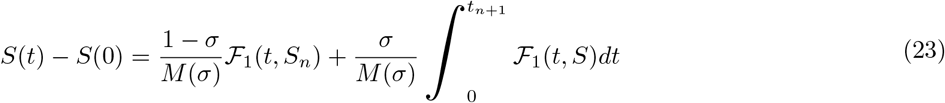

The successive terms is then given below:

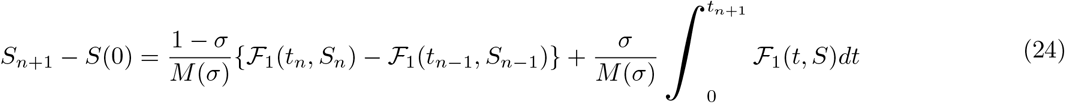

For a closed interval [*t*_*k*_, *t*_(*k*+1)_], the function ℱ_1_(*t, S*) can be interpolate by the interpolation polynomial

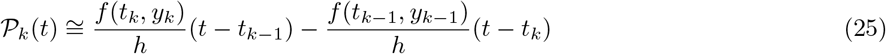

for *h* = *t*_*n*_ − *t*_*n*−_. Calculating the integral in (24) by using equation (25) we have that

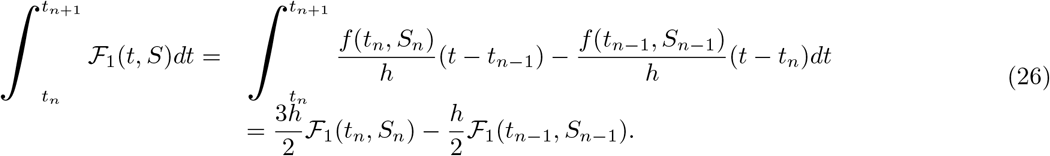

Together with (24) and (26) and after some algebraic simplification we get

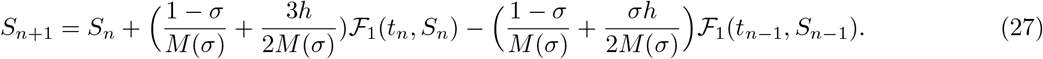

Similarly, the recursive formulae for the rest equations of of system (2) is given as follows

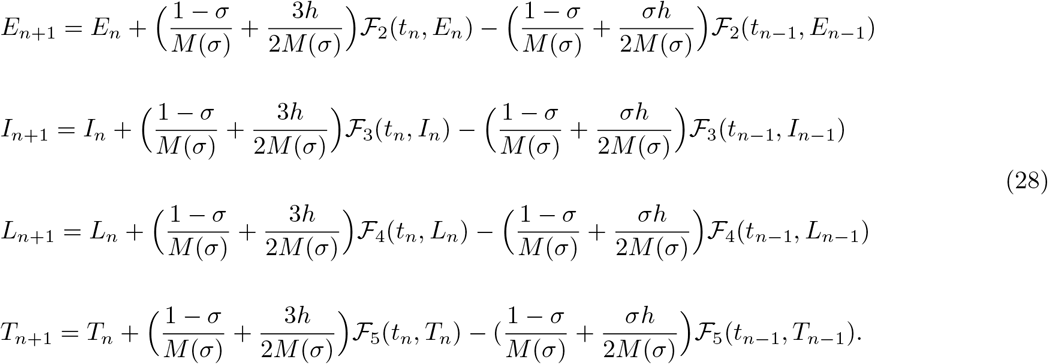

## 5. Syphilis model with ABC operator

Applying the definitions of ABC as in [30] to model (1) we have the following system of equations

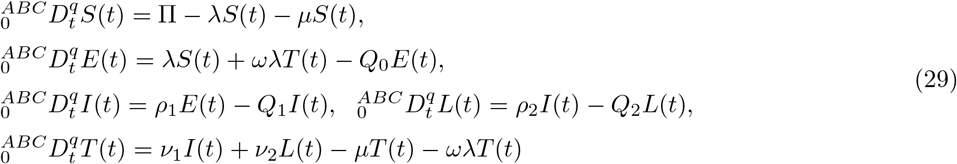

where *q* is the fractional order, subject to initial conditions

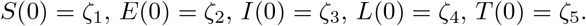

### 5.1. Existence and uniquness of solutions of the ABC model

Here, we use the fixed point theory to show the existence and uniqueness of the solutions of the system (29). consider the ABC system (29) re-written in the form below

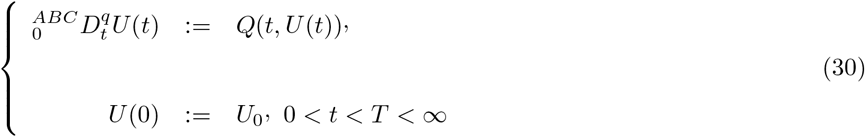

where *U* (*t*) = (*S, E, I, L, T*) and *Q* is therefore a continuous vector function given as

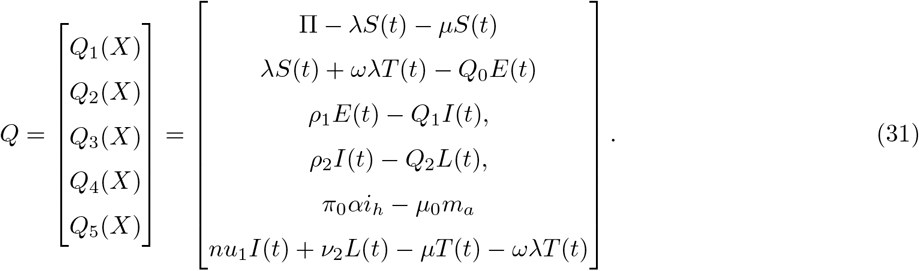

and *U*_0_(*t*) = *S*(0), *E*(0), *I*(0), *L*(0), *T* (0) represents the initial conditions of the state variables respectively. The function *Q* satisfies the condition for the Lipschitz continuity and can be described as

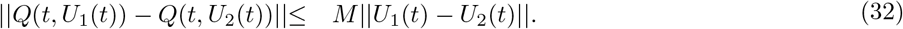

We give the following result, to show the existence and uniqueness of model (29).

#### Theorem 8.

Existence and uniqueness

The ABC model given by (29) has a unique solution if the following condition is satisfied

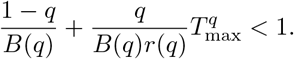

*Proof*. Applying the AB-fractional integral on the system (30), we obtain the following non-linear Voltera integral equation:

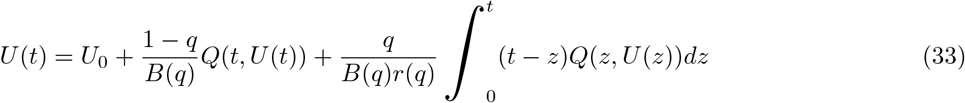

We assume that *J* = (0, *T*) and consider the operator *ϕ* : *C*(*J*, ℝ^5^) → *C*(*J*, ℝ^5^), defined by

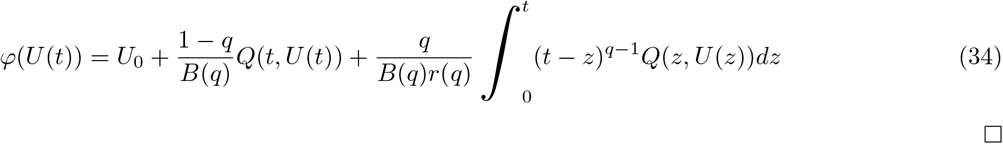

□

Equation (33) becomes

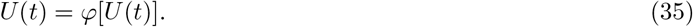

The supremum of *J*, ||.||_*J*_ is

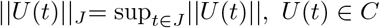

where *C*(*J*, ℝ^5^) along the norm ||.||_*J*_ represents a Banach space. Also,

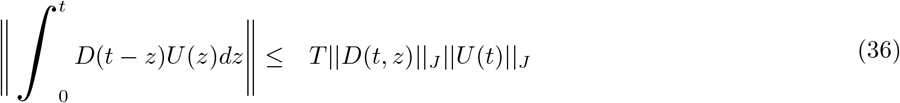

with *U* (*t*) ∈ *C*(*J*, ℝ^5^), *D*(*t, z*) ∈ *C*(*J*^2^, ℝ^5^) such that

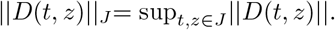

Applying the definition of *ϕ* as stated in (35), we have

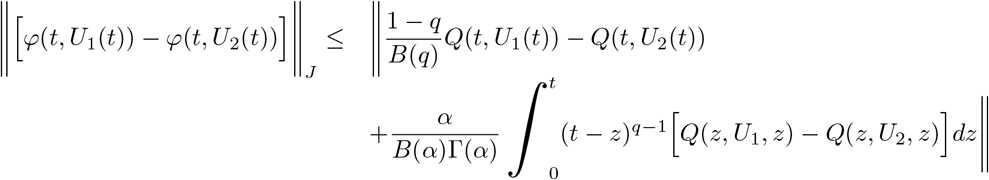

Using the Lipschitz condition stated in (32) coupled with the result obtained in (36) and the principle of triangular inequality, we get the following after some algebraic manipulations

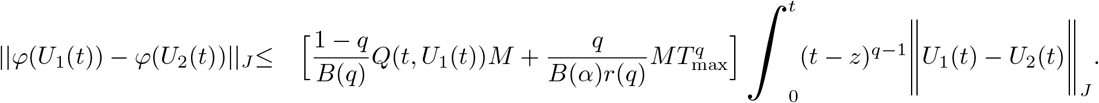

We thus have

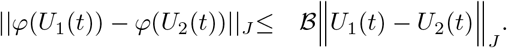

where

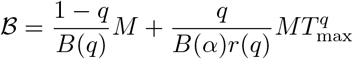

Therefore, the operator *ϕ* will become a contraction if the condition (32) holds on *C*(*J*, ℝ^5^). By the Banach fixed point theorem, the system (31) has a unique solution.

### 5.2. Numerical scheme for the ABC model

In this subsection, we derive the numerical scheme of the syphilis model in (29) using the Adams-Bashforth method [31]. Consider the system (30) written as follows

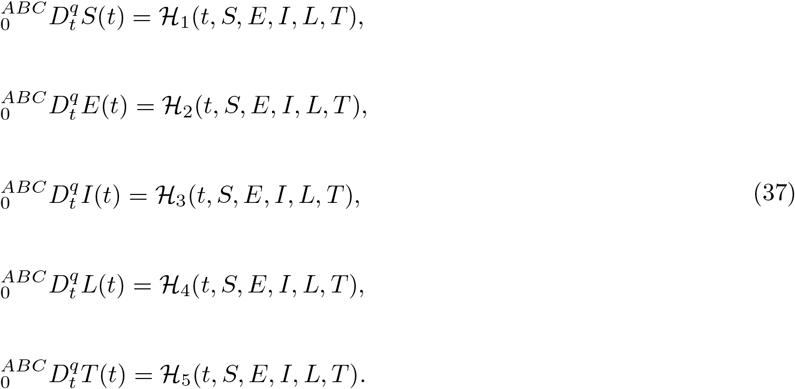

Using the fundamental theorem of fractional calculus, the system (37) is converted to the following

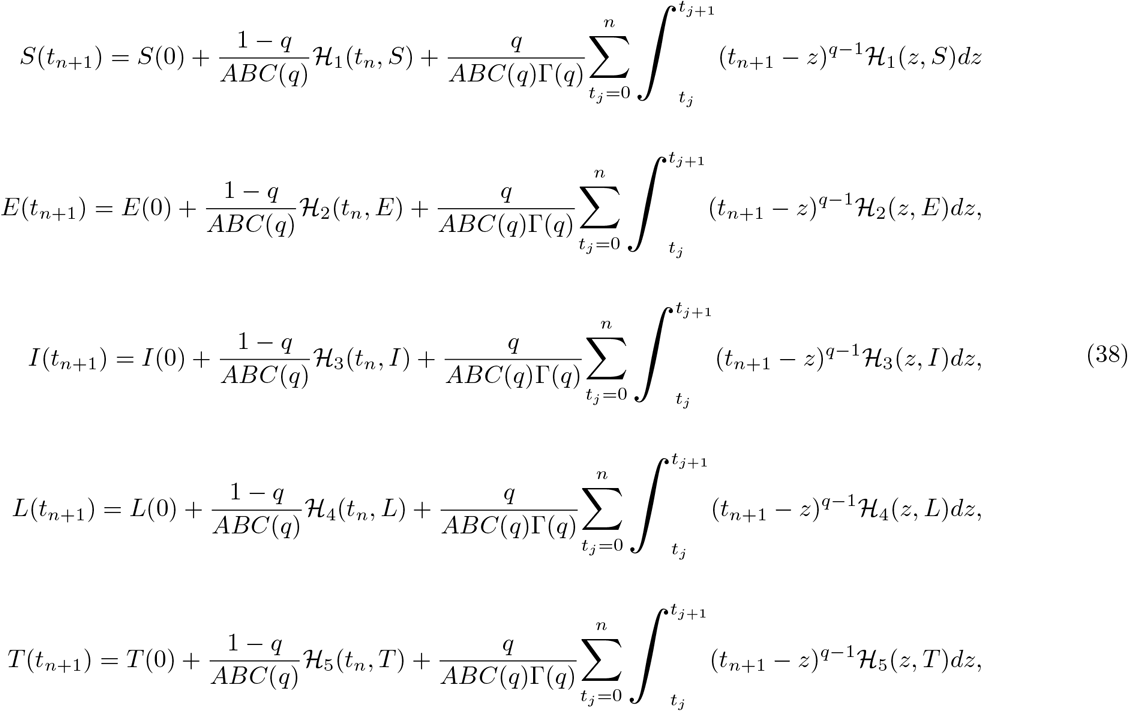

Note that the integrals in (38) are approximated through the two-point interpolation polynomial, hence we have the iterative scheme for the Syphilis model (29). After some algebraic calculations, we obtain an approximate solution for the ABC Syphilis model as follows

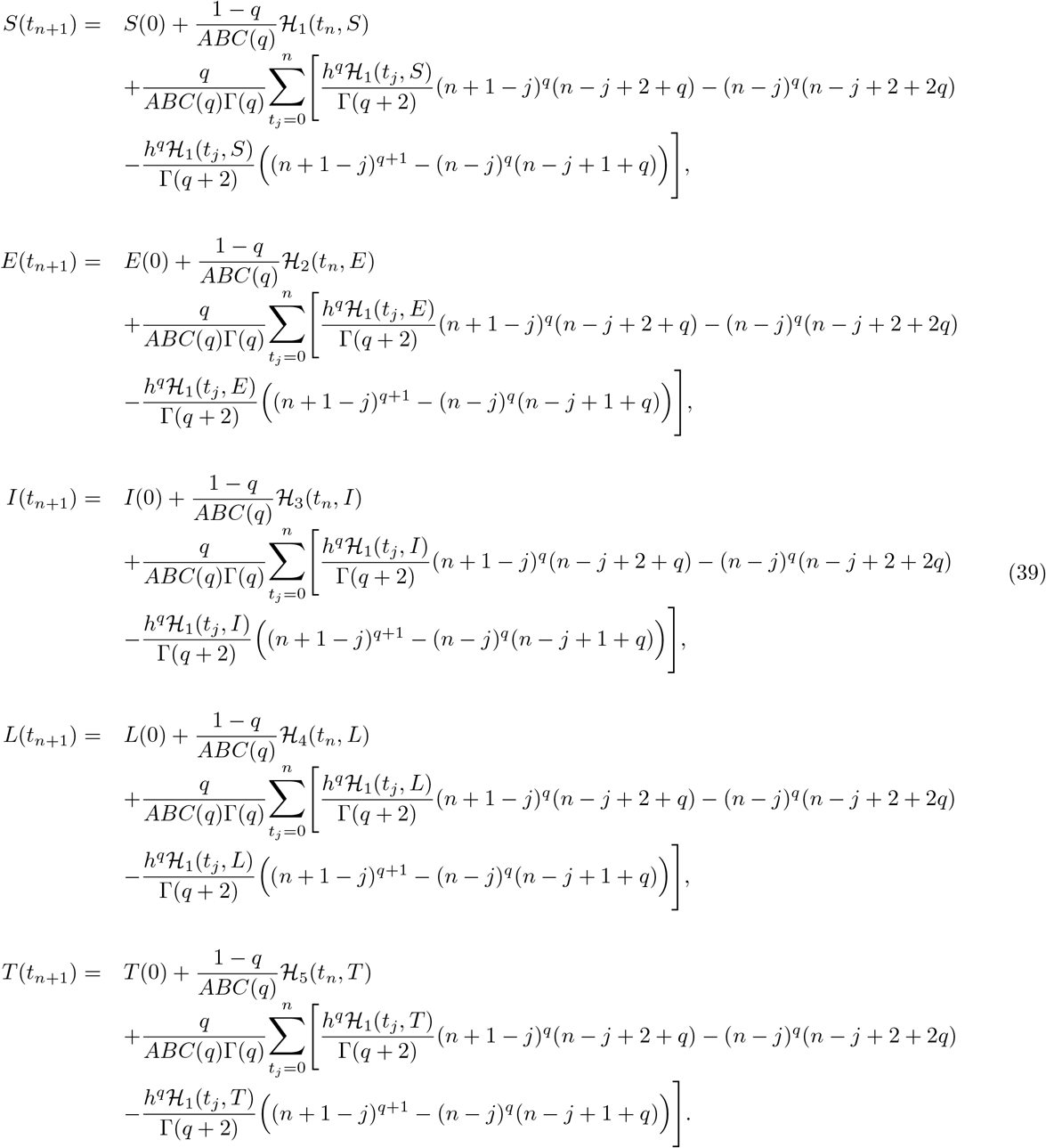

Next, we use the numerical scheme to simulate the results of the ABC Syphilis model.

## 6. Numerical simulation

In this section, we explored the numerical dynamics of Syphilis model (2) in the context of Caputo-Fabrizio. In this work, the following parameter values and initial conditions were used

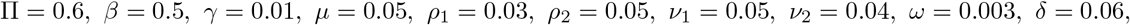

with units per day and the initial conditions *S*(0) = 14, *E*(0) = 2 *I*(0) = 0 *L*(0) = 0 and *T* (0) = 4 for our numerical simulations.

Figure 2(a) is the susceptible individuals (*S*) for integer and non-integer. The number of susceptible humans reduces as the fractional order *σ* derivatives increases from 0.3 within 120 days. Figure 2(b) is the exposed humans (*E*) in which both integer and non-integer order derivatives presented. As the fractional order derivative increases from 0.3, the number of exposed humans increased. This is naturally expected as more humans are exposed to the disease. Figure 2(c), the number of infected individuals at early stage (*I*) increase as the fractional order derivatives reduces from 0.9 to 0.3. This is naturally the case as more humans move into infected humans compartment. Figure 2(d) is the individuals at late stage infection with syphilis and as the fractions, order derivatives increases from 0.3 to 1, the individuals increase towards the non-integer. In Figure 2(e), the number of individuals treated reduces as the fractional order derivatives decrease. In effect, to increase the number of individuals treated, the fractional order would be reduced to 0.3.

**Figure 2:**
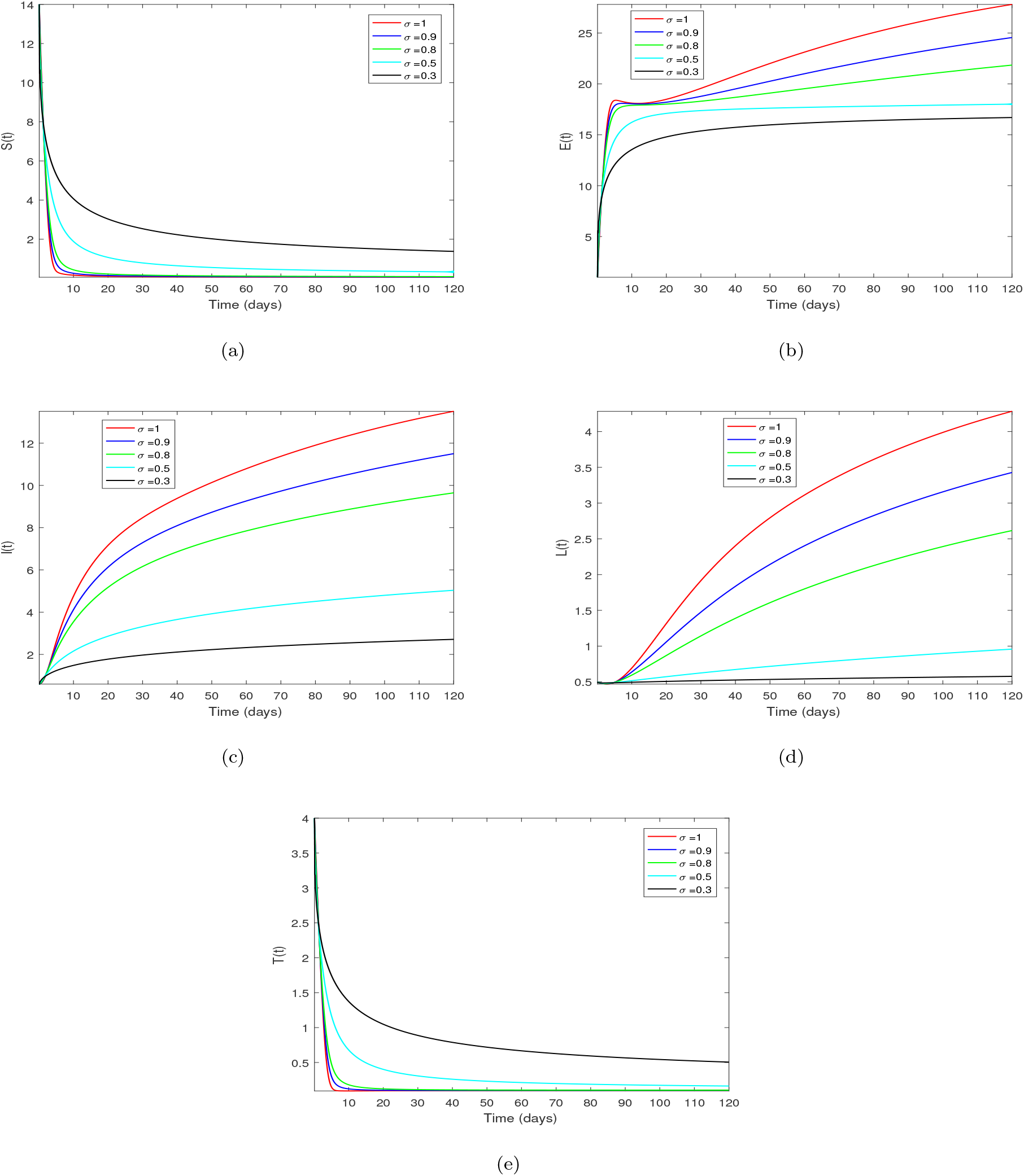
Simulations for Syphilis model (2) for the susceptibles, exposed, infected at early stage, treated and individuals at late stage of Syphilis via Exponential-law at *σ* = 1, 0.9, 0.8, 0.5, 0.3.

Figure 3 was obtained by solving system equation (29) with Mittag-Leffler function using the numerical scheme of equation (39). The same parameter values and associated initial conditions were used for this work simulations. Figure 3(a) is the susceptible individuals (*S*) and as fractional order derivative increases, the number of susceptible reduces. Figure 3(b) depicts the exposed individuals (*E*) in which the number of exposed individuals (*E*) increase as the integer order 1 turn to non-integers. Figure 3(c) shows that the early infected individuals (*I*) with syphilis increases as the non-integer fractional order derivatives 0.3 increases toward the integer order 1. The Figure 3(d) is the late individuals infected with syphilis (*L*) in the community where fractional order derivatives increase from 0.3 upward towards integer order 1. Figure 3(e) is the treated individuals (*T*) where the non-integer order derivatives increase from 0.3 towards 1, the number of individuals under treatment reduce.

**Figure 3:**
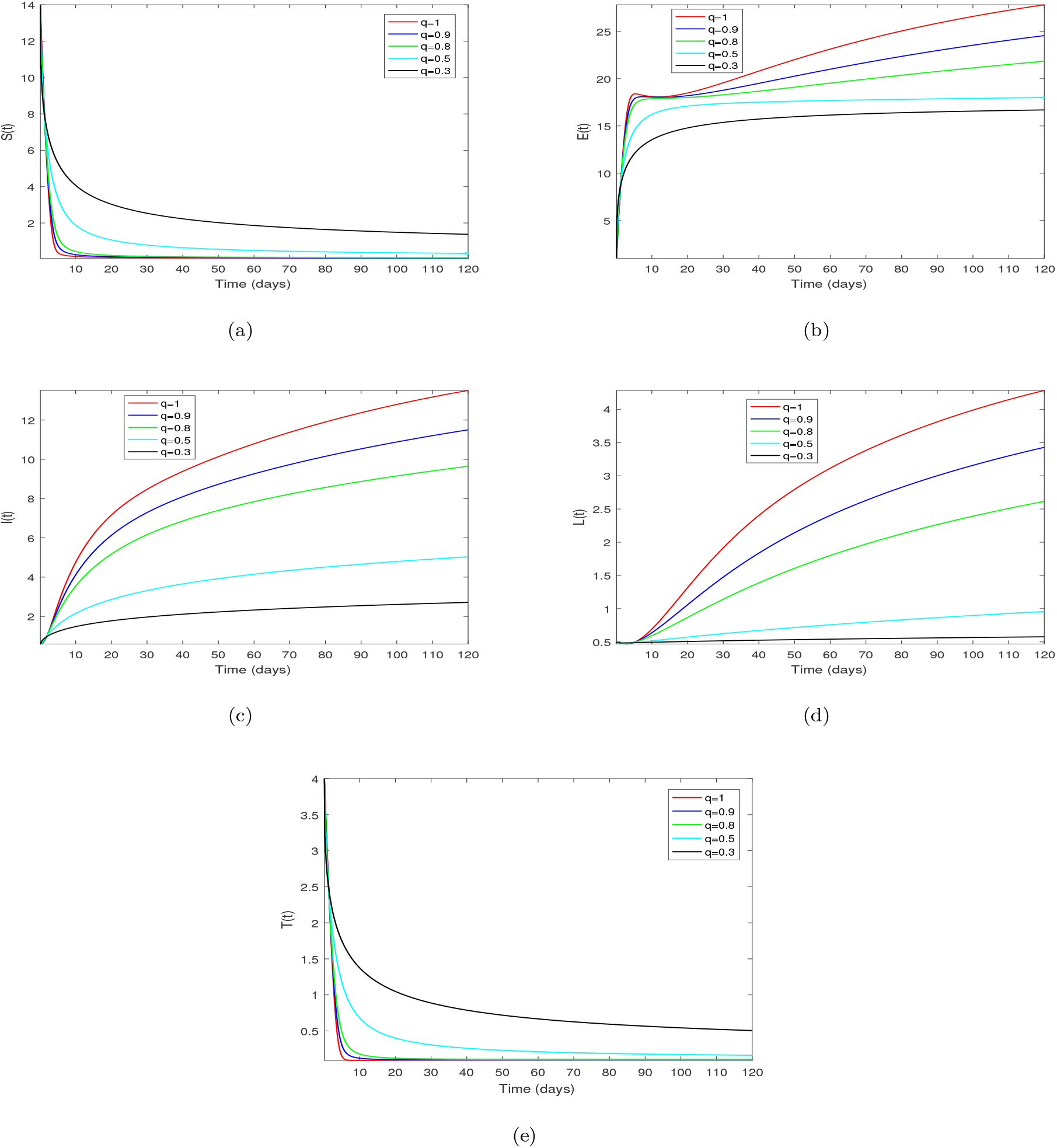
Simulations for Syphilis model (29) for the susceptibles, exposed, infected at early stage, treated and individuals at late stage of Syphilis via Mittag-Leffler function at *q* = 1, 0.9, 0.8, 0.5, 0.3.

## 7. Conclusion

In this study, a syphilis mathematical model with Caputo-Fabrizio and Mittag-Leffler function was formulated and analysed. The basic properties of the model were examined and the steady states of the model were investigated. The stability analysis of the disease free was carried out and found to be stable from both local and global perspectives. In each operator used for the study, the existence and uniqueness of solutions was established. Respective numerical schemes for each operator was carried to obtain numerical simulation to support the analytical solution. It was established that the fractional order derivatives influence the dynamics of the Syphilis disease in the community. It is suggested other complex models can be investigated using fractional derivatives operators.

## Data Availability

None

## Acknowledgements

The authors will like to say thanks to their respective Universities for the production of this manuscript.

